# Domain specific need assessment for Hospital Disaster Preparedness: a systematic review and critical interpretative synthesis

**DOI:** 10.1101/2022.04.04.22273416

**Authors:** Neeraj Pawar, Raghvendra Gumashta, Girjesh Gupta, Rajendra Mahore, Jyotsna Gumashta

## Abstract

**Background:** During disasters, the most pressing demands are those related to health, and hospital preparedness is an area that require special attention. Hospitals are viewed as resources that must be proactively utilized in the event of a disaster. If national and local systems, particularly health systems, are unprepared to deal with disasters, the vulnerability of both individuals and communities is amplified. The unexpected surge in demand for important health services caused by disasters frequently overwhelms health systems and institutions, leaving them unable to perform the life-saving measures that are required. This study aims to understand various domains of hospital disaster preparedness by critically synthesizing qualitative evidence from selected research on the topic.

**Methods:** Electronic data base from PubMed, Google Scholar, key hospital disaster related journals was explored with search syntax focusing on hospital related disaster preparedness. Peer reviewed English articles published from January 2011 were systematically selected and critical interpretative qualitative synthesis was done to have comprehensive understanding of the said phenomenon.

**Results:** A total of 29 articles were included in the systematic review. Major resultant domains describing disaster preparedness were Human Resource, Logistics And Finance, Response, Communication, Coordination, Patient Care, Evacuation and Personal Protection. Some domains were more emphasised than others, this information can help prioritizing the action based on need especially in the times of disaster.

**Conclusion:** Disaster preparedness needs a comprehensive approach including context specific optimization with the effective use of available resources.

## 1. Introduction

Disaster is defined as an incident or condition that exceeds the local community’s ability to manage the impact. It is a major deterioration of society’s function at any degree posed by hazardous occurrences combined with situations of exposure, vulnerability, and capacity, resulting in one or more of the following losses and impacts: human, material, financial, and environmental. Natural disasters have wreaked havoc throughout history. The disaster’s impact can be rapid and confined, but it’s usually extensive and can persist for a long time. The effect may put a community’s or society’s ability to cope with its resources to the test, necessitating aid from other sources, such as national or international levels. (1)

As one of the most significant healthcare institutions, hospitals play a critical role in providing services in both routine and disaster-struck situations. (2–4) Hospitals are considered safe and sophisticated places for patient care and must ideally be sufficiently equipped to respond to a variety of foreseeable and unpredictable incidents. (5–7) Despite their reputation, hospitals are vulnerable to both internal and external calamities. (8) Intrinsic disasters like biological emergencies, fire, earthquakes damage the hospital structure and function, compromising the delivery of essential care services and potentially posing threat to the health of hospital residents, including the staff, leading to the facility’s entire evacuation. (4)

According to the World Health Organization (WHO), disaster preparedness includes identifying potential threats, building an inventory of existing assets, and establishing a capacity to respond to a health crisis. When it comes to hospital-specific emergency actions, the WHO defines disaster preparedness as an intersectoral collaborative effort to respond to, control, and recover from an imminent hazardous event. (9) The Sendai Framework provides more detailed definitions of disaster preparedness, including disaster drills and the establishment of support networks. (10)

Hospital-related disaster preparedness warrants a holistic approach with the inclusion of structural building plans that has provisions for fire safety, earthquake resilience, inbuilt management system, trained human resource, logistics, communication, and transport mechanisms. There is a spectrum of research published on hospital-related disasters, some are learning experiences from incident-specific hospital-related disasters, others include the impact of training, qualitative inquiry, meta-analysis, all discussing the key aspects of hospital-related disaster-specific preventions and interventions. (11–35) This brings a fragmented yet comprehensive knowledge base on the various aspects of disaster preparedness. This study, therefore, aims to generate a qualitative pool of ideas from the existing body of literature, by observing and reporting the emphasized areas /domains in individual studies and contextualizing the evidence presented to have an overall understanding of the needs of hospital disaster preparedness and resilience.

## 2. Methods

### 2.1 Adopting a novel approach

The current research conducted a systematic review with critical interpretive synthesis. This design is based on qualitative research processes incorporated in traditional systematic review methods, and it can be used to combine qualitative and quantitative evidence. It is mainly geared toward hypothesis generation with the provision of a recursive and adaptive approach to searching for literature and selecting materials for inclusion in reviews. It treats the literature as deserving of critical examination in and of itself by analysing its underlying assumptions; and achieves synthesis through a logical balance between evidence and theory. The emphasis is not only on the narrative context but also on the ‘authorial voice’ for evidence generation. (36)

### 2.2 Identifying relevant studies

The Preferred Reporting Items for Systematic Reviews and Meta-Analyses (PRISMA) standards were followed for conducting this review. (37), see S1File, S2 File.

Figure 1 is a flow chart depicting the selection process. A search strategy was created based on the PRISMA protocol, and screening, study selection, quality evaluation, and data extraction were all completed.

**Figure 1:**
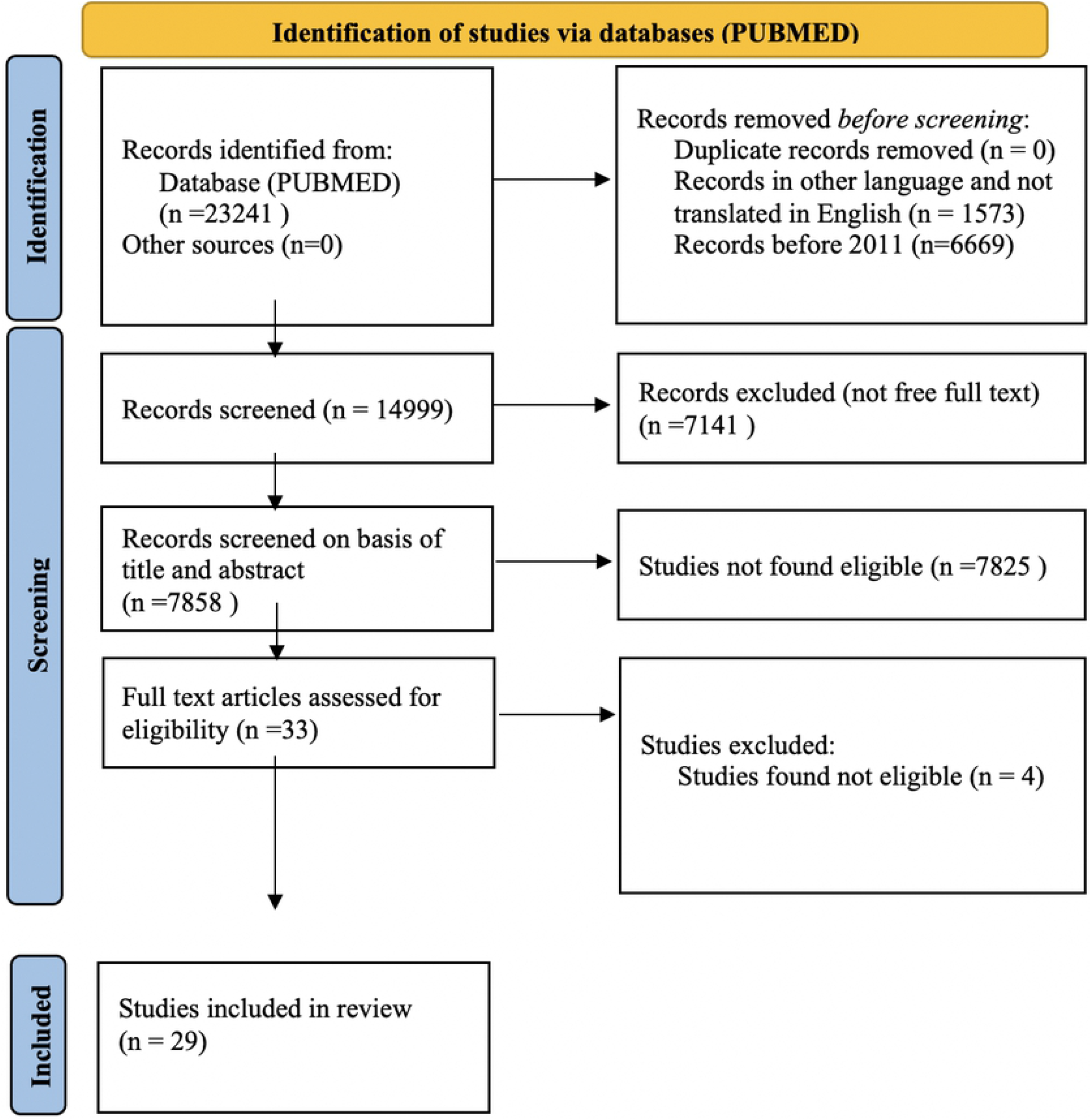
PRISMA flow diagram for new systematic reviews which included searches of databases.

Data sources used in this study included MEDLINE (PubMed), Google Scholar, key journals such as Disaster Medicine and Public Health Preparedness, BMC health services and research, PLOS ONE, Journal of Education and Health Promotion, and reference lists of selected articles and systematic reviews. Keywords were selected, based on relevance to the study context, consultation with scientific experts, and terms used in the relevant literature as a guide. “Emergency Evacuation”, “Urgent Evacuation”, “Evacuation Time”, “Patient Evacuation”, “Medical Facility”, “Health Center”, “Healthcare Center”, “Tertiary Referral Center”, “Tertiary Care Center”, “Hospital”, “Disaster”, “Event”, “Incident”, “Emergency”, “Hazard”, “Fire” were among the keywords used in this study. The initial search syntax was first developed for PubMed, employing operators, keywords, and search fields. Studies completed between January 2011 and December 2021, were included in the literature search. Table 1 shows the search method that was used.

**Table 1:**
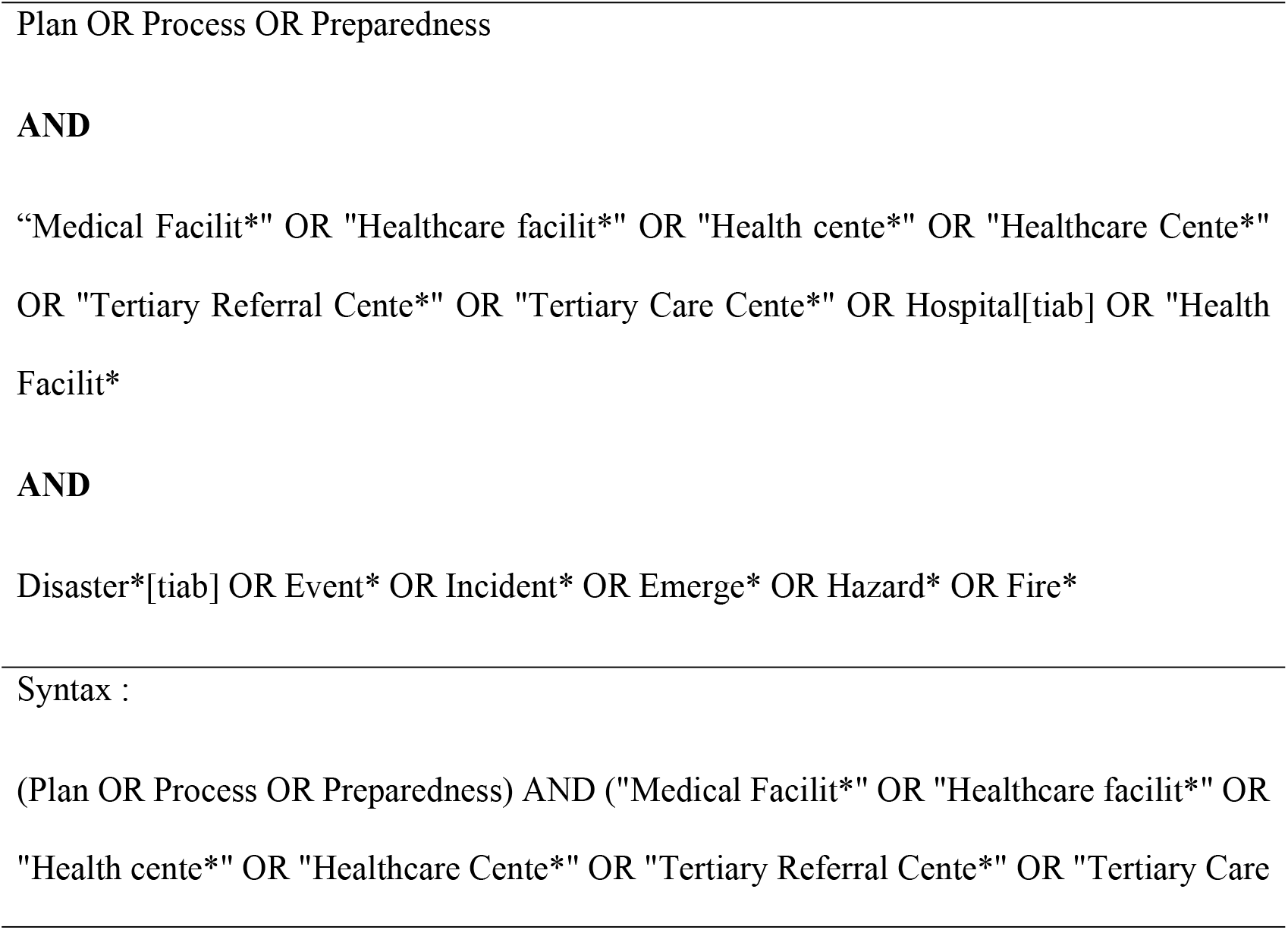
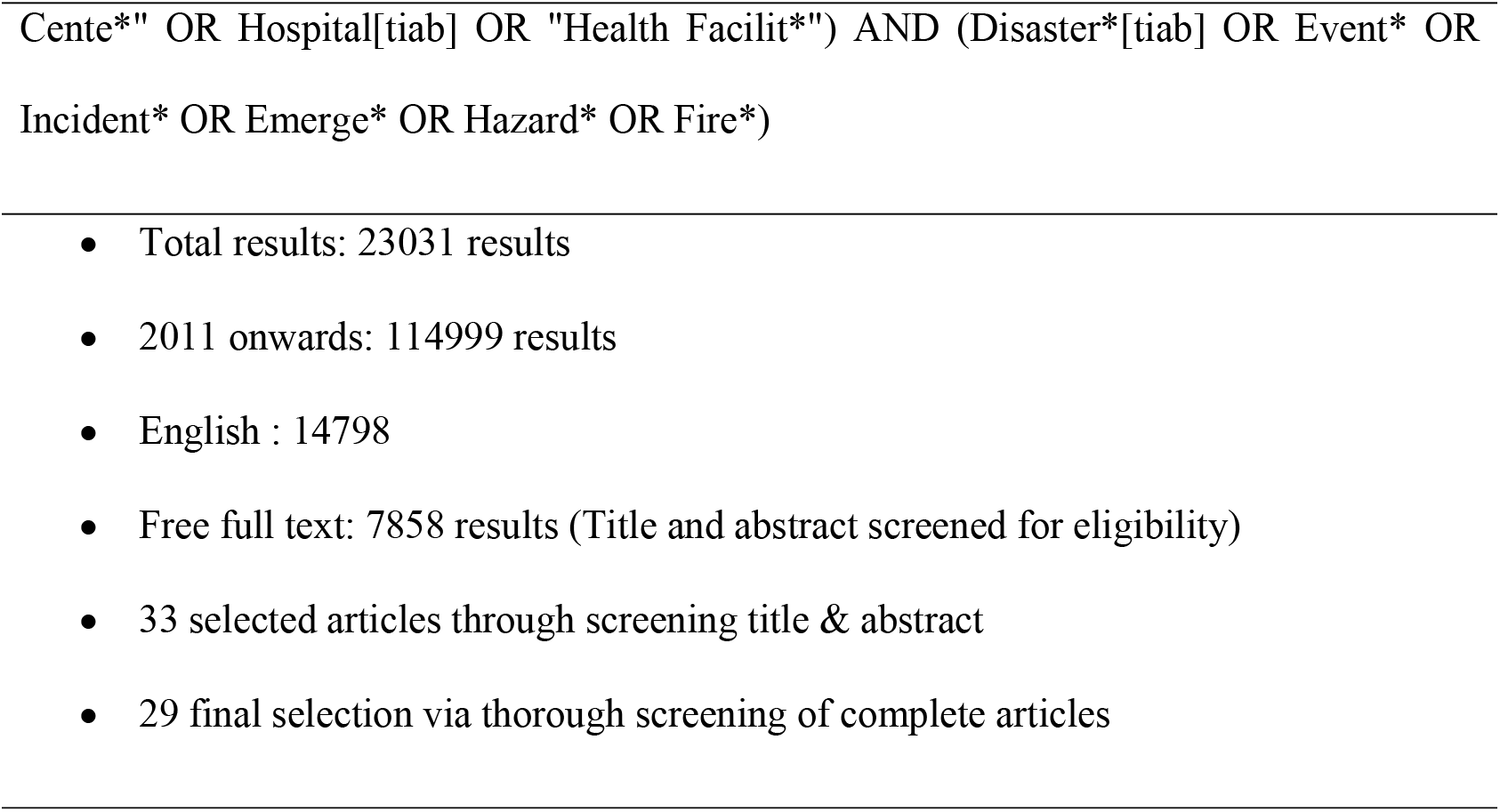
Search strategy terms.

### 2.3 Study selection

After applying this search syntax, a total of 23241 records were obtained out of 7858 articles were selected for screening after applying relevant filters, like removal of duplicates, records in other languages, and records before 2011. Then in the next step, the titles and abstracts were screened for eligibility. (Figure 1) For evaluating the quality of papers, to be included, rather than focusing on specific study categories or publications that satisfied specific methodological standards, we attempted to prioritize studies that looked to be relevant. Two researchers worked independently on study screening and selection, as well as data extraction. We considered it was critical to use a low barrier for an interpretive review to maximize the inclusion and contribution of a wide range of papers at the concept level. As a result, we took a two-pronged approach to quality control. First, we opted to eliminate publications that were shown to be grossly inconsistent. Second, as we’ll see later, once in the review, the synthesis itself required critical judgments and interpretations of acceptability and contribution. Therefore, appraisal prompts for informing judgments about the quality of papers were also used which included the following questions:

1. Are the research’s goals and objectives described clearly?
2. Is the research methodology defined and relevant for the study’s goals and objectives?
3. Do the researchers provide a detailed overview of how their findings were replicated?
4. Is there enough evidence to back up the researchers’ interpretations and conclusions?
5. Is the analytical procedure appropriate and well-explained?
6. In the end, the review included a total of 29 papers.

### 2.4 Summarizing the selected studies

Based on the aforementioned criteria, two researchers (RG and NP) extracted the final study data separately. A summary of the selected studies, including the names of the initial authors, year, location, design, and the key findings are reflected in Table - 2. Among the 29 selected studies, two were systematic reviews with meta-analyses, one was a qualitative study, rest were descriptive studies. (Table-2)

**Table 2 :** Overview of studies included in the review after systematic selection and the extracted key words from the articles on the factors affecting disaster preparedness of in hospital setting.

| Codes | First Author<br>,Year | Place | Type | Method | Number of<br>Reference | The findings from the<br>articles |
| --- | --- | --- | --- | --- | --- | --- |
| **A** | Sellers (39),<br>2021 | Austra<br>lia | Review<br>Article | Concept<br>Analysis | 30* | Effective Utilization Of<br>Space, Increased<br>Demand Of Logistics,<br>Human Resource,<br>Training And Call Back<br>Mechanism |
| **B** | Sahebi (42),<br>2021 | Iran | Original<br>Article | Qualitati<br>ve | 56* | Response To The<br>Incident, Hospital<br>Preparedness, Moral And<br>Ethical Values, And<br>Human And<br>Environmental Factors<br>Affect The Process Of<br>Emergency Evacuation. |
| **C** | Shin (41),<br>2021 | Korea | Retrospecti<br>ve Study | Descripti<br>ve | 34* | Mitigation, Preparedness<br>, Response ,Recovery |
| **D** | Wood (40),<br>2021 | Iraq | Review<br>Article | Review | 46 | Fire Safety And Hospital<br>Management, Fire Safety<br>Response, Emergency<br>Risk Management |
| **E** | Woyessa<br>(11), 2020 | Ethio<br>pia | Original<br>Article | Cross-Sectional | 32* | Planning, Committee,<br>Human Resource,<br>Logistics Finances,<br>Communication |
| **F** | Bazyar (12),<br>2020 | Iran | Review<br>Article | Systemat<br>ic<br>Review<br>With<br>Meta-Analysis | 50* | Conducting Exercise,<br>Training Program, Proper<br>Management, Effective<br>Use Of Resources, |
| **G** | Goniewicz<br>(13), 2020 | Polan<br>d | Review<br>Article | Retrospe<br>ctive<br>Analysis | 42 | Emergency Evacuation<br>Plan, Evacuation Drills,<br>Specific Responsibility,<br>Frequent Checks |
| **H** | Jang (14),<br>2020 | South<br>Korea | Retrospecti<br>ve study | Retrospe<br>ctive<br>Data<br>Analysis | 7 | Triage, Full Utilization<br>Of Resources, Medical<br>Staff Safety |
| **I** | Farcas (15),<br>2020 | USA | Model<br>Developme<br>nt | Model<br>Develop<br>ment | 17* | Communication,<br>Resource Use, Staff And<br>Patient Safety, And<br>Maintenance Of Roles |
| **J** | Bhattacharya<br>(16), 2019 | India | Original<br>Article | Intervent<br>ion<br>Study | 17 | Emergency Plan,<br>Disaster Committee And<br>Personnel |
| **K** | Beyramijam<br>(17), 2019 | Iran | Original<br>Article | Intervent<br>ion<br>Study | 27* | Planning, Triage,<br>Education, Space Usage,<br>Human Resource |
| **L** | Kazemzadeh<br>(18), 2019 | Iran | Review<br>Article | Systemat<br>ic<br>Review<br>With<br>Meta-Analysis | 22 | Plan , Policy, Inadequate<br>Resources, Guidelines |
| **M** | Wax (19),<br>2019 | Canad<br>a | Review<br>Article | Review | 24 | All Hazard Versus<br>Hazard Specific Plan,<br>Specific Roles,<br>Interprofessional Efforts |
| **N** | Lee (20),<br>2018 | China | Original<br>Article | RCT | 37 | Training, Evacuation<br>Procedure, Fire Drills,<br>Self-Rescue, Patient<br>Evacuation |
| **O** | Koka (21),<br>2018 | Tanza<br>nia | Original<br>Article | Cross-Sectional | 34 | Disaster Plan ,Training,<br>Triage, Communication<br>System , Human<br>Resource , Space |
| **P** | Shalhoub<br>(22) , 2017 | Saudi<br>Arabi<br>a | Original<br>Article | Cross-Sectional | 16 | Disaster Plan,<br>Committee, Exercises<br>,Staff, Transport, Surge<br>Capacity |
| **Q** | Tekin (23),<br>2017 | Turke<br>y | Review<br>Article | Review | 29 | Communication, Role Of<br>Media, Humanitarian<br>Services, Triage,<br>Disaster Plan |
| **R** | Lakbala (24),<br>2016 | Iran | Original<br>Article | Cross-Sectional | 18 | Limited Beds, Limited<br>Human Resource, Drills,<br>Training, Space |
| **S** | Paganini<br>(25), 2016 | Italy | Original<br>Article | Cross-Sectional | 29 | Preparedness , Training,<br>Triage ,Activate And<br>Deactivate Plan |
| **T** | Ingrassia<br>(26), 2016 | Italy | Original<br>Article | Cross-Sectional | 40 | Command System, Surge<br>Capacity, Simulations &<br>Drills , Recovery Plan |
| **U** | Lofqvist (27),<br>2016 | Swed<br>en | Original<br>Article | Cross-Sectional | 18 | Written Plan, Education<br>& Training, Specific<br>Plans, Practice Over<br>Planning |
| **V** | Zhong (28),<br>2014 | China | Original<br>Article | Cross-Sectional | 58 | Essential Services,<br>Cooperative System,<br>Incident Command<br>System, Specific Plan |
| **W** | Djalali (29),<br>2014 | Europ<br>e | Original<br>Article | Cross-Sectional | 19 | Education And Training,<br>All Hazard Approach,<br>Multisectoral Financing,<br>Collective Strategic Plan |
| **X** | Talati (30),<br>2014 | India | Manual | Manual<br>Drafting | 11 | Disaster Committee,<br>Mock Drills, Command<br>Centre, Training,<br>Communication, Family<br>And Media Handling |
| **Y** | Djalali (31),<br>2013 | Swed<br>en | Original<br>Article | Cross-Sectional | 38 | Contingency Plan,<br>Committee , Logistics,<br>Finances |
| **Z** | Femino (32),<br>2013 | USA | Review<br>Article | Retrospe<br>ctive<br>Review | 7 | Communication, Onsite<br>Triage, Drill, External<br>Partners |
| **AA** | Sobhani (33),<br>2012 | Iran | Original<br>Article | Cross-Sectional | 25 | Communication,<br>Logistics, Administration<br>, Infrastructure |
| **AB** | Venkatasesha<br>n (34), 2011 | India | Review<br>Article | Narrative<br>Review | 8 | Emergency Evacuation,<br>Fire Drill , Staff<br>Education, Space, Sign<br>Post |
| **AC** | Murphy (35),<br>2011 | United<br>Kingdom | Original<br>Article | Cross-Sectional | 15 | Unit Design, Equipment,<br>Planning, Training<br>Triage |

**\*references includes systematic review &/RCTs**

The collected data were analysed using critical interpretive analysis. At the outset, a review question was formulated with the scope of modification. Initially, one author (NP) read the entire text of each of the 29 articles that met the criteria. All of the codes and basic concepts linked to disaster preparedness aspects were retrieved for coding. Then they were examined line by line, numerous times, to get the beginning codes. Following that, the first two authors (RG, NP) compared and contrasted all the identified codes, and then grouped related codes into one category to generate a sub-theme.

### 2.5 Synthesis of argument

The goal of the analysis was to provide a critically informed integration of information from all of the articles in the review. The synthesized argument was then linked to a pre-existing structure, the WHO Hospital Safety Index Evaluation Form - Module 4 (38), to help formulate the major themes. Finally, all the researchers reviewed the summarised texts to generate new ideas, and the findings including the synthesized themes and subthemes were appropriately elucidated and mapped. Analysis was done in MS office 365, and graphical representation including geo-mapping was done using Tableau version 2021.4. Necessary modifications were made until everybody concurred on the final version.

## 3. Results

To understand the geo-spatial prominence of selected studies in terms of disaster preparedness and its aspects, included studies were numbered and geo-mapped, and average scores for each country on the domain-wise emphasis were assessed on the Likert scale (no, moderate, prominent emphasis). Computed scores were added at a country level, a reverse red color gradient of which is reflected in the map with 10 (Dark red) being the lowest scores for Australia and 78 (Light red) being the heights average score for Ethiopia. (Figure 2)

**Figure 2:**
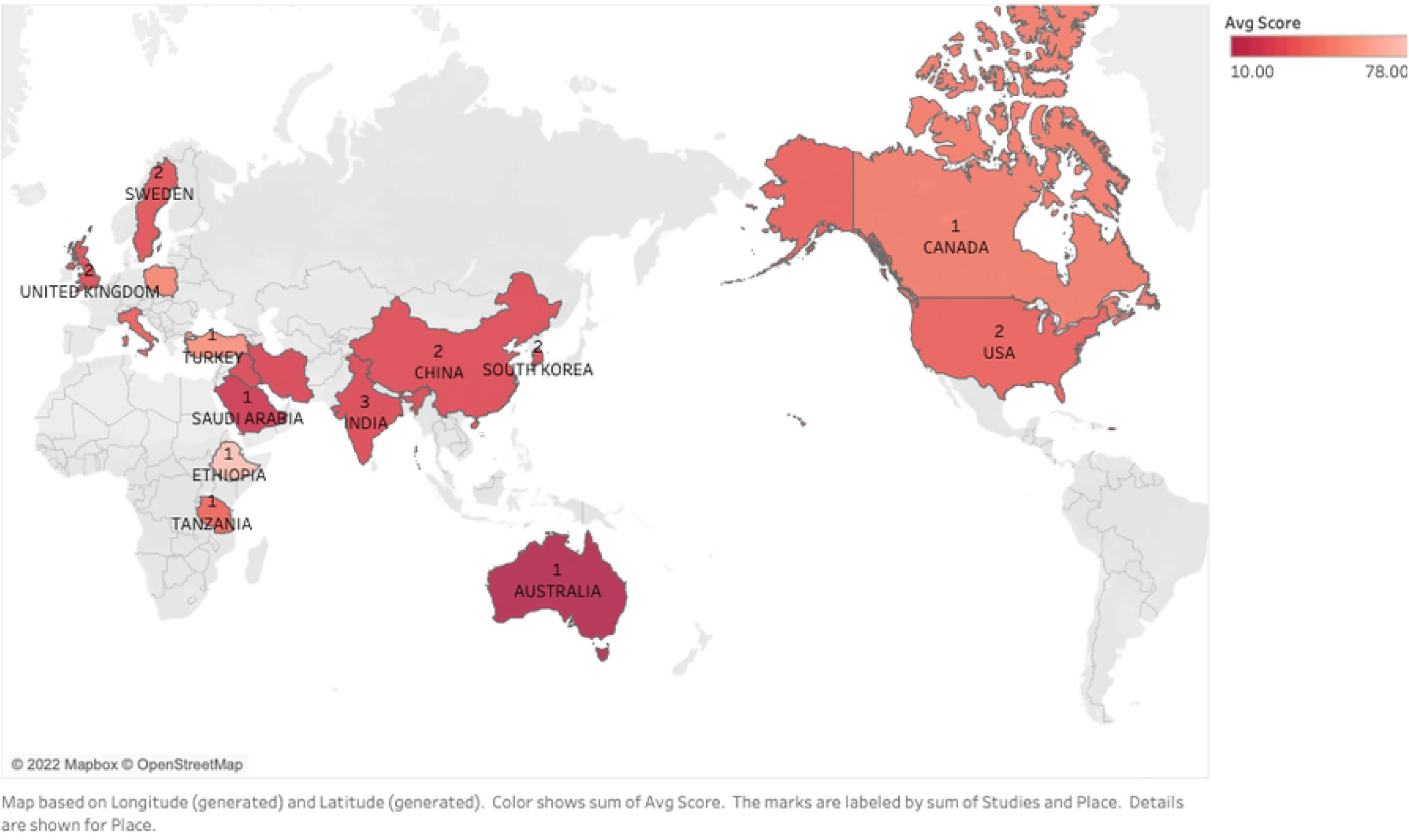
World map showing number of included studies from different countries and country-wise gradient of average scores on disaster preparedness.

In this, each study was also evaluated based on core domains of disaster preparedness derived from the WHO Hospital Safety Index Evaluation Form. The evaluation was done by qualitatively analysing each article with a special focus on the authorial voice including typographical emphasis (Italics usage for keywords, stage cues, backstories, and capitalization) and contextual perspectives. The emphasis of each disaster preparedness domain in the individual study is mapped using a heat map with the color orange reflecting no emphasis (coded as 0), yellow with moderate emphasis (coded as 1), and green reflecting prominent emphasis (coded as 2). Included studies presented on x axis with their code reference (A-AC) are mentioned in Table 2 and disaster preparedness domains are reflected on the right side in tabular form. (Figure 3). A study conducted in Ethiopia ticked most boxes (reflected in green) for prominent emphasis as it covered all key aspects of disaster preparedness prominently featuring in the background statement, study tools, discussion, and conclusion. (Figure 3)

**Figure 3:**
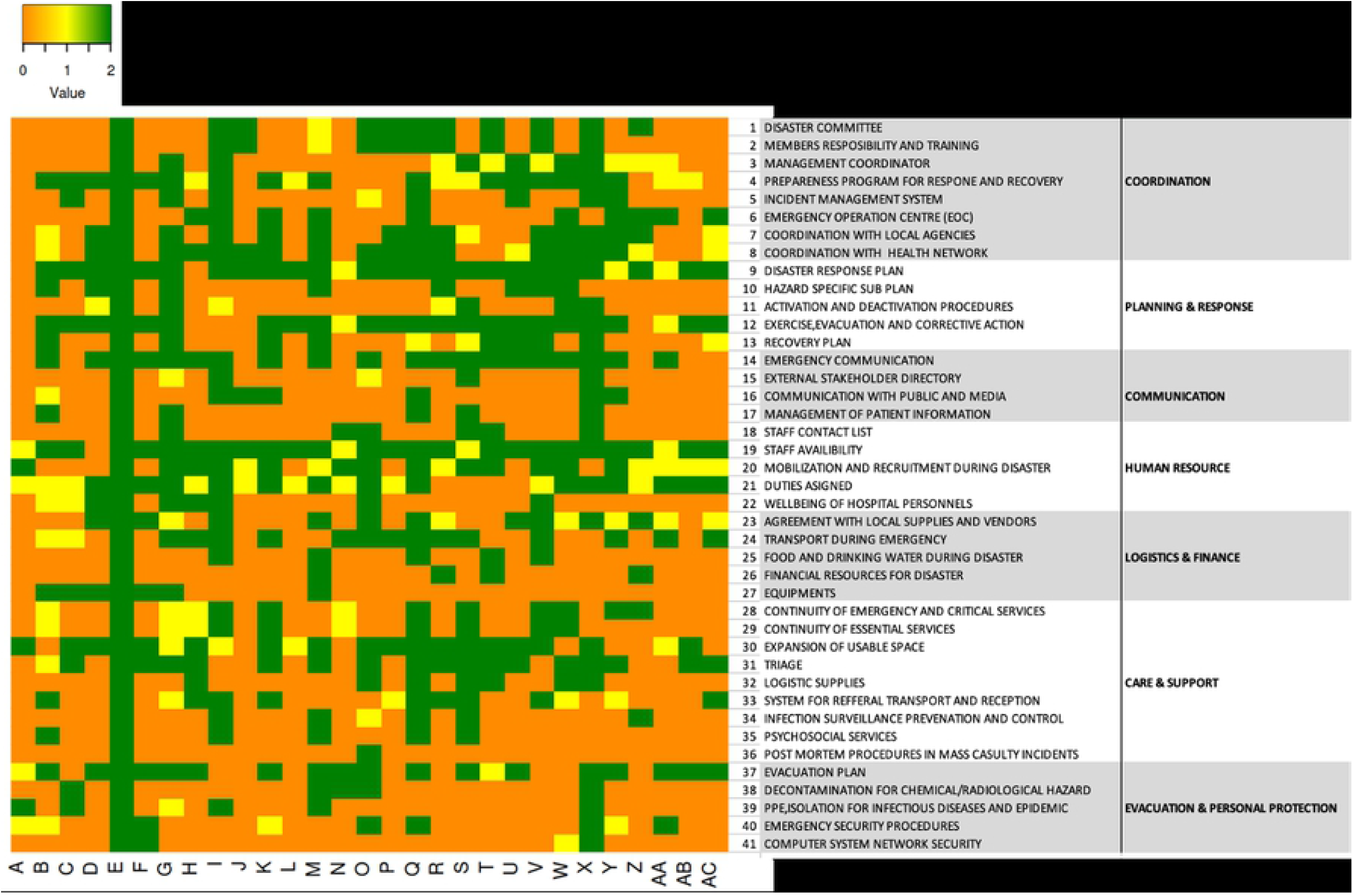
Heat map showing individual study focused emphasis on attributes of disaster preparedness in the included studies.

The magnitude of reasoned representation of key focused areas in included studies is illustrated through a word cloud. In this, the commonly repeated keywords and concepts from each study were compiled to understand the ictus and weightage of various disaster preparedness components. As it is reflected some of the key insistent words were “disaster plan”, “human resource”, “equipment and logistics”, “triage”, “drills and triage”, “command system”. (Figure 4)

**Figure 4:**
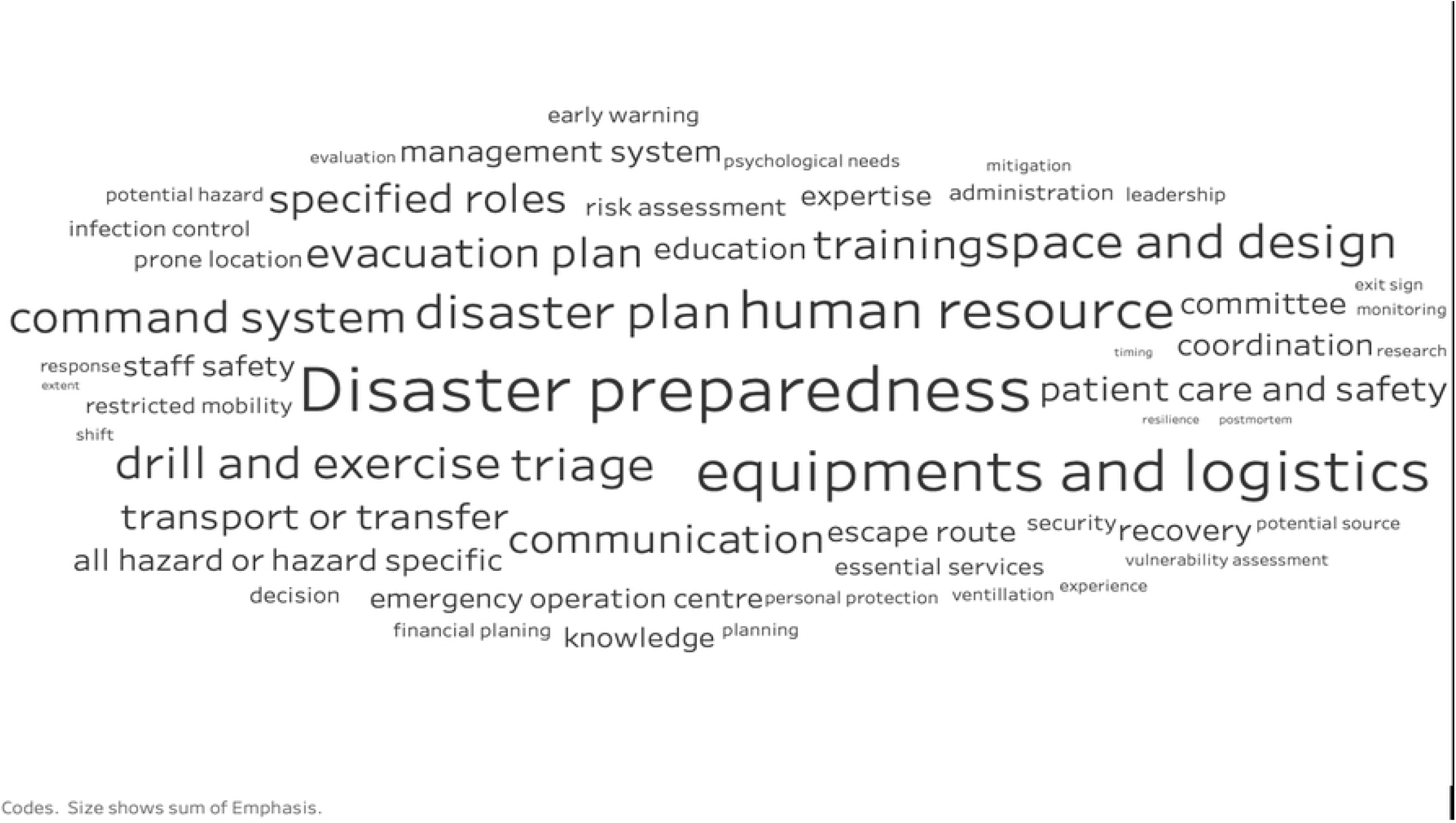
Word cloud depicting reasoned and proportionate representation of key focus areas of the shortlisted studies irrespective of their geographical representation.

Based on the results obtained from the word cloud, and heat map the clarity of purpose and strategic priorities in various important domains and subdomains of disaster preparedness was jotted and themed and domain-wise emphasis scores were graded and computed, then the proportions were mapped as individual slices of nested pie diagram. As reflected, Human Resource, Coordination, Logistics And Finance, Response, Patient Care and Support, Communication, Evacuation, and Personal Protection emerged as major themes and among subthemes: staff availability, specified roles, evacuation plan, disaster response plan, exercise/drills, internal and external communication, local supplies were the prominent ones. (Figure 5)

**Figure 5:**
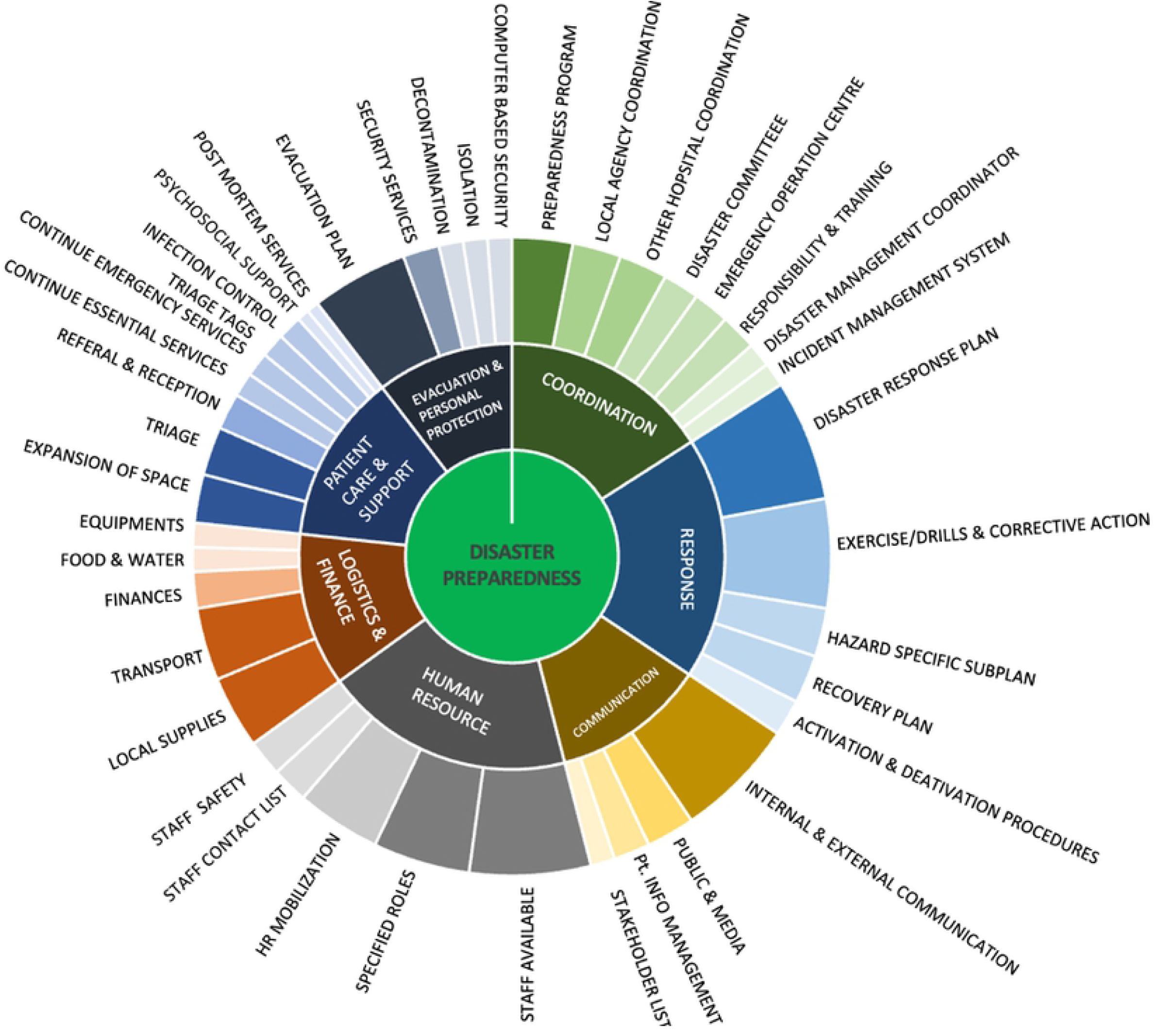
Nested pie showing domain specific emphasis of disaster preparedness in the included studies.

## 4. Discussion

Hospital preparedness for disasters differs significantly from other area of disaster preparedness. Although papers detailing hospital disaster preparedness are common, most studies are incident-specific and they are unable to give a thoroughgoing insight into all the elements of the all-hazard disaster preparedness approach. Also, systematic reviews done on the topic quantified the evidence from individual studies leading to a telescopic perspective and have the potential to miss some key ideas and dynamics related to disaster preparedness. This study uses a novel qualitative interpretative synthesis to not only understand the hospital preparedness dynamic in the context of disaster but also help generate new insight making the scope of vision broader and thus helping understand the concept better. We here discuss key elements emphasized in the included studies.

### Prioritizing intervention based on risk perception

A concept analysis conducted at the Griffith University of Australia emphasizes using a comprehensive solution-based strategy for the best possible service provisions in addition to addressing the needs for training, retraining, capacity building, and mock drills. The effect of changing modalities and processes of existing healthcare facilities requires scrutiny at all levels of needs and demands for making the facility friendly to a high influx of people struck by disaster. When one attribute is extended beyond normal operational capacities, the effectiveness and capacity of the other attributes will likely be limited. (39)

The safe functioning of electrical equipment prevents fire hazards through strategic preparedness for any accident, emergency, and unforeseen circumstances at various health care service facilities. The overall perspective hence should be focused upon consolidation and matching of risk perception, related factors, and assessment-based prioritized reforms cum rapid corrective actions. (40) The inverse proportion of single attribute preponderance with other related attributes has been emphasized in a study guide to set up a balanced environment of facilities and manpower to address the overwhelming load for treatment care and support in situations of disaster occurrence with or without warning. (39)

In an exclusive study conducted in Gumi City, the Republic of Korea, the issues identified therein include conceptual clarity for preparedness, mitigation, response and rehabilitation, and reconstruction cyclically through detailed analysis of requirements in a hospital setting for minimizing the risk through assessment of manpower, skill, knowledge, experience, expertise, vulnerability, capacity, and inbuilt systems towards disaster response. There has been stress on the felt need of having an overall control network for cohesive actions by the administration, management, and facility (41)

### Comprehensive planned response

The protocol development, amendment, and timely up-gradation for disaster management plans as per demographic needs requires sustained and sustainable inputs, efforts, and actions (25) A study conducted in Iran has categorically highlighted, evidence for considering the importance of an all-inclusive response consisting of preplanning, execution of a plan, and information dissemination through a systematic approach. The areas identified by the researchers include mobility, flexibility, resonance, interaction, speed, accuracy, awareness, respectability, positivism, and spirit of faith among service providers and beneficiaries. (42)

European countries have stretched upon evidence-based fulfilment of technical requirements to ensure safe, secure convenient, methodological appropriate, and cost-effective evacuation measures supported by a well-devised plan of action with pre-decided and pre-identified roles and responsibilities of various stakeholders using the resources available including but not limited to the aspects of feasibility, speed, and accuracy of evacuation exercises while using interactive mechanisms of security among government organizations, Non-government agencies and service providers. (13)

Planning has been identified as the key driver of change as it reflects the policies and defines the directionality of guidelines and procedures to be adopted for adequate resource utilization and contingency plan based implementation of preventive processes under the umbrella cover of accreditation standards universally accepted for response to disasters in various departments of the hospitals. (18) Facility-based preparedness may not be in tune with the resource mobilization cum appropriate measures undertaken at the national level and it, therefore, necessitate the need for developing regional, local, and facility-specific plans under the umbrella cover of national health policies, priorities, and planned public health preferences. (26)

An actionable strategic plan helps in clarifying the readiness related to the operational plan, contingency plan, and resource plan in emergencies while considering the magnitude and strength of the expected hazards. The hospital disaster plan hence is dependent on the dual edge of the economy and dedicated planned inputs. (31) Planning is not only essential for getting the personnel educated and trained but also for ensuring methodical approaches towards the inclusion of micro-plans related to evacuation, triage, and communication. (32)

### Prioritized and categorized qualitative

Development of the domain-specific surveillance system and establishment of resource strengthening measures aid in designing the comprehensive action plan for hospital disaster resilience. It should define and describe the role of emergency medicine response, staff capability, capacity building, communication systems, interventional plans, recovery and rehabilitation, logistics management, and standardized practices cum networking. The categorization of disaster management-related supplies has also a pivotal role in addressing the basic needs of food transport medical supplies and preventive measures along with the availability of common medications. (28)

There is hence, a transformational requirement for the inclusion of thus identified capacity enhancement measures into policy and programmatic documents to enable the ready availability, uniformity, and methodical approach of these measures at the implementation levels in various health care facilities. (42)

The administrative, managerial, and technical collaboration between the gas industry and hospitals is essential for appropriately, justifiably, and efficiently merging the objectives of patient safety at levels of awareness of hazards, capacity building, communication, monitoring the equipment and their maintenance. (40)

Having established a web-based disaster response, and mitigation system with well-defined processes using available resources along with structural reforms for an easy, smooth and clear flow of information, communication, and operations will do wonders for logistically managed and research-supported mechanisms of operation for disaster preparedness. Given this, there has to be a resonating balance between the existing health system network and model-based command and control systems. (15)

It is also understood through various studies that setting up of an intrinsic web of interactive, mutually sportive, and vibrant communicative link will enable the sharing of preventive measures among the facilities, especially in isolated hospital disasters. However without the political will and visionary support of all stakeholders the visibility, preference, and outcome of the interventional public health design may be compromised or even paralyzed during the actual occurrence of disasters. Area demarcation, command and control, and recovery mechanisms are to be defined assessed, monitored and evaluated on a regular predefined timeframe and set norms for such practices. (26)

Categorization of any hospital preparedness for various levels of disaster risk is found dependent majorly upon human resources. It has been found that there may be a mismatch of risk perceptions by the administration and the actual scenario at facilities, and hence there is underlined the categorical need for a comprehensive interventional setup supported by regular assessment, monitoring, and evaluation. (11) The emphasized areas generally include evacuation preferences, communication, logistics, human resources, commanding, and management. It is a common phenomenon observed during the disaster to have a surge in the affected population, making facilities insufficient to cater for the requirements to be addressed as per globally set standards for indicators of health facility preparedness. (12)

Multisectoral, multidimensional, multifunctional, and multiple team-based designs of disaster prevention and vulnerability reduction exercises enable the hospitals to address the hazard, area, and mechanism-specific stakeholder participation through disaster exercises distributed among various subunits of the hospital systems inclusive of diagnosis, treatment, care, and support. (19)

### Health system strengthening

Preparedness of the hospitals averts the deaths and prevents complications among the survivors as the risks are minimized through effective interventions of vital health care services. There is, however, a thin line of demarcation between structural and non-structural safety as both of these overlap each other’s objectives. (12)

A participatory approach for devising, defining, and developing the comprehensive list of responsibility distribution, disaster manual development, and greater in-depth understanding at all levels of health care functionaries ensures adequacy in resources capabilities and preparedness for minimizing the risks, threats, and vulnerabilities related to hospital disasters. (30) Incident command systems equipped with modern gadgets like CCTV and cameras help ensure effective administrative managerial and communication systems. These also guide for assessing the training needs of individuals and groups as per their area wise interventional requirements. (33)

### Decision tree approach based tools

Detailed analysis of situations, existing policies, structure, equipment, command system, planning, and monitoring based systems are the need of the hour for adequate, appropriate and effective alignment of priorities in line with the available resources motivational status of implementors, and defined networks of response. (35)

Strengthening of health systems depends primarily on the capacity of facilities service providers and mechanisms for strengthening the training status in an ongoing updated manner to ensure optimal gains in assurance for disaster preparedness. (29) The analysis of strengths, weaknesses, opportunities and the threat of disaster risk based on improvement requirements for hospital readiness are extensively analysed for improvement in the action plans and programs of disaster management. The internal weakness identified in studies has been a lack of exit signs, and a weak telecommunication system whereas the strengthening factors have been trained manpower, travel support mechanisms, and improvement in the infrastructure of the hospital. The opportunities for improvement are immense including facilities for shifting, emergency support for health care, and spaces for the affected population, whereas population density and disaster-prone houses are threats during the disaster. (24)

The training of health care workers especially those involved in those areas having vulnerability to fire hazards have been recommended using awareness sessions generated enhanced knowledge and action-based for expected readiness of quick response to have effective actions towards fire prevention, rescue and evacuation exercises. (20)

Hospital disaster preparedness depends not only upon the sharing of the information, awareness generation, and technical capabilities but also depends on the training, re-training, disaster drills, research, and adherence to the emergency disaster plans. (22)

It has been underlined that the capacity building for disaster preparedness, management, and administrative infrastructural support shall go a long way in establishing a self-sustainable model for the action-oriented, automated, and competent response to hospital disasters of varied nature as evidenced during the mass causalities and distribution of workloads of frontline divisions of critical care facilities. (25)

The deficiencies identified in addressing disasters include non-translation of written protocols into practice, in coordination among the various agencies, capacity building lapses, and non-observance of repeated practical sessions and surprise alarm system based check exercises for emergency evacuation, saving lives, shifting patients, and hassle-free transit systems. (27)

Detailed decision trees have been charted for the evacuation and its triage with a support system through field hospitals providing emergency medical, humanitarian, and logistics facilities.(23) Triage providers without any formal training may be influenced by mass casualties and therefore biased support to select individuals or groups belonging to influential classes although not falling in the priority list of triage categorization. It, therefore, necessitates the emphasis on in-depth understanding, training, and assistance of reliable communication networks with tools, systems, and warning mechanisms in place. (21)

Triage has been identified as a key instrument during disasters of various nature affecting countries. The classification of triage into categories of severe mild and dead with their respective color coding and related prioritization cum transportation with an inherent limitation for follow-up of casualties on a long-term basis, it becomes difficult to assess the outcome of mass casualty and to understate the effect of preventive interventions in place for medical assistance. (14)

Development and implementation of established, modern, and new tools for prevention and control of hospital disasters include careful observance of well-displayed evacuation routes, evacuation areas, and related medical care facilities with equal emphasis on training of all identified personnel.(34) Operational research is required for the assessment of interventional requirements so as to develop specific need-based interventions with assisted audio-visual support through mock drills, seminars, and educational interventional packages. (16)

The use of the ‘Hospital Emergency Response Checklist’ is found to be very useful for policy development, decision-making regarding response plans, training at various levels, and reaching conclusions based on the results obtained through risk and hazard assessment. There is a need to conduct studies related to a multidimensional assessment of the effect of the intervention, especially in the hospitals identified previously as having a low level of disaster preparedness. (17)

### Strength and Limitations

This research comprehensively reviewed, analysed, and discussed all possible facets of disaster preparedness using a fairly unprecedented qualitative approach that adds strength to the research. The review search includes articles published in the English language, and those indexed in PubMed, Google Scholar, and some key selected journals on disaster management, so articles from other databases (including grey literature), and studies from different languages on disaster preparedness could have been potentially missed. To overcome this issue, we used a robust and exhaustive search syntax, with the provision of all possible synonyms of keywords, in PubMed, allowing initial screening of the vast majority of articles.

## Conclusion

The present study concludes that multisectoral initiatives are key to the formulation of successful hospital disaster preparedness. Monitoring of this preparedness plan includes all important ingredients related to governance, logistics, program management specific need-based arrangements, use of new tools, efficient communication systems, modernization of health care facilities, and well-defined triage systems in place.

## Future scope

This study can act as a guiding paper for the policy makers and program managers not only to recognize the key focused areas in disaster resilience but also to understand the dynamics of disaster management, helping them adapt to the emerging needs of hospital disaster preparedness. The ideas generated in the present study, mark a stepping stone in the directionality of developing a comprehensive, all-inclusive disaster resilience framework and formulation of universal tool of intervention in future for different geo-social settings.

## Data Availability

All relevant data are within the manuscript.The study is a qualitative synthesis from selected studies.

## Ethical considerations

Ethical issues (Including plagiarism, misconduct, data fabrication and/or falsification, double publication and/or submission, redundancy, etc.) have been completely observed by the primary author.

## Notes

### Competing Interest Statement

The authors have declared no competing interest.

### Funding Statement

The author(s) received no specific funding for this work.

### Author Declarations

The study is a systematic review with critical interpretative synthesis and no ethical issues are involved

